# A Machine Learning Classification of Individuals with Mild Cognitive Impairment into Variants from Writing

**DOI:** 10.1101/2024.02.16.24302965

**Authors:** Hana Kim, Argye Hillis, Charalambos Themistocleous

## Abstract

**Introduction:** Individuals with Mild Cognitive Impairment (MCI), a transitional stage between cognitively healthy aging and dementia, are characterized by subtle neurocognitive changes. Clinically, they can be grouped into two main variants, namely into patients with amnestic MCI (aMCI) and non-amnestic MCI (naMCI). The distinction of the two variants is known to be clinically significant as they exhibit different progression rates to dementia. However, it has been particularly challenging to classify the two variants robustly. Recent research indicates that linguistic changes may manifest as one of the early indicators of pathology. Therefore, we focused on MCI’s discourse-level writing samples in this study. We hypothesized that a written picture description task can provide information that can be used as an ecological, cost-effective classification system between the two variants.

**Methods:** We included one hundred sixty-nine individuals diagnosed with either aMCI or naMCI who received neurophysiological evaluations in addition to a short-written picture description task. Natural Language Processing (NLP) and BERT pre-trained Language Models were utilized to analyze the writing samples.

**Results:** We showed that the written picture description task provided 90% overall classification accuracy for the best classification models, which performs better than cognitive measures.

**Discussion:** Written discourses analyzed the AI models can automatically assess individuals with aMCI and naMCI and facilitate diagnosis, prognosis, therapy planning, and evaluation.

## 1 Background

With the growth in the number of older adults, age-related neurodegenerative diseases such as Alzheimer’s disease (AD) have dramatically increased. These neurodegenerative diseases cause a great deal of financial and emotional burdens not only for patients and their caregivers but also for society. The global cost of dementia care was estimated to exceed $500 billion in the United States [1]. It is expected to rise to $2 trillion by 2030 [2]. Research has suggested that the preclinical phase of dementia may start earlier than the diagnosis. Detecting the preclinical stage of dementia and providing an intervention will delay the onset of AD. This will significantly minimize the socio-economic burden, which is expected to reduce societal costs by 40% [3].

Mild cognitive impairment (MCI) is an intermediate stage between cognitively healthy aging and dementia [4]. It represents a critical preclinical stage of the AD [5–7]. MCI includes four different clinical subtypes. Two main subtypes are amnestic MCI (aMCI) and non-amnestic MCI (naMCI); this subtyping is determined based on the impairment in memory. Individuals with aMCI are characterized by memory loss, while individuals with naMCI demonstrate impairment in domains such as executive functions, attention, and language [8, 9]. Also, depending on the number of cognitive domains impaired, individuals can be categorized into single-domain and multi-domain MCI. Although a higher risk of developing dementia characterizes individuals with MCI, not all individuals with MCI will progress to dementia; some may remain stable, and others even regress to a condition of healthy aging [10–12]. Therefore, it is essential to discriminate against those who are more likely to progress to dementia for early intervention since most treatment strategies are more effective in the presymptomatic stage of dementia [13].

Depending on the two main subtypes of MCI, differences in the progression from MCI to dementia have been reported. In general, it has been suggested that aMCI represents the earliest symptomatic manifestation of AD pathophysiology, while naMCI is likely to progress to non-Alzheimer’s dementia [14–16]. A recent 20-year retrospective study supports this and adds more information with a large dataset (N = 1188). The authors demonstrated that aMCI represents a greater risk for progressing to dementia (not only for AD) compared to naMCI. The odd ratio of the progression to dementia between aMCI and naMCI was statistically different [17]. This highlights the clinical need of a robust, reliable system for classifying aMCI and naMCI [18].

There have been several approaches for MCI diagnosis. Behaviorally, a brief cognitive screening test can assist in identifying whether an individual has an apparent cognitive impairment [9]. Neuropsychological tests can be administered depending on the need for further assessments to determine the presence or degree of impairment in cognitive functions. The tests for MCI biomarkers require magnetic resonance imaging (MRI) or lumbar puncture for cerebrospinal fluid (CSF). Increased amyloid burden was found to be specific to aMCI, while naMCI does not exhibit a specific abnormality in neuroimaging (see review for Yeung, Chau [19]). Blood biomarkers, considered a comparatively more straightforward means of testing, have also been investigated [20]. Unfortunately, such tests for MCI biomarkers are not routine care in clinical settings [21–24]. Moreover, the cost and availability of the testing technique (e.g., MRI) may limit its impact on individuals’ care [25].

Linguistic changes are considered to manifest as one of the earlier indicators of pathology in cognitive impairment. It has been reported that they emerge years before deficits in other cognitive systems become apparent [26]. In particular, writing is a cognitively and linguistically complicated activity. Writing consists of distinct phases: planning, generating, and revising [27]. Writers initially set a goal for organizing their knowledge and executing the plan in response to the topic of the writing activity. Then, writers revisit and revise their output. All phases should be well orchestrated to accomplish successful writing within cognitive systems such as executive functions, attention, and working memory. A recent review article highlighted the diagnostic value of writing tests, especially at the discourse level (Kim et al., 2023). Discourse is any language beyond the sentence level [28, 29]. Kim and colleagues (2022) investigated the prognostic value of discourse-level writing tests. They conducted a chart review of individuals diagnosed with MCI and visited a neurology outpatient clinic more than once (N = 71). They classified the study participants into the stable MCI group and the converter group. The authors examined whether a written discourse task using the Cookie Theft picture [30] predicts clinical course in the MCI group. They found that the stable MCI group produced more core words than the converter group at their baseline assessment. This underscores the potential clinical utility of discourse-level writing tasks for early detection of those who are likely to progress to dementia from MCI.

In recent years, computational methods such as Natural Language Processing (NLP) have been used to analyze written language samples in individuals with neurodegenerative disease [31–36]. Computational methods offer two advantages. First, they allow the elicitation and combination of measures from different linguistic domains. A decisive property of ML models is their ability to find patterns between features associated with a specific group of individuals, i.e., patients with aMCI and naMCI.

Earlier studies successfully distinguished healthy adults from individuals with MCI from healthy adults [32], MCI from dementia [37–40], and the subtypes of primary progressive aphasia [41, 42]. These findings highlight the role of ML as an important method that can contribute to the existing approaches [35] and to inform clinical assessment and therapy.

This is the first attempt to classify two subtypes of MCI (aMCI vs naMCI) using discourse-level writing samples in NLP. Since writing involves several cognitive functions (especially language, vision, and motor control), we hypothesized that a written picture description task could distinguish individuals with aMCI and naMCI. This work could potentially provide a quick and easy tool to facilitate diagnosis from written language tasks.

## 2 Methods

### 2.1 Participants

Our participants were comprised of 169 individuals diagnosed with either aMCI or naMCI. All individuals were recruited through the Johns Hopkins Hospital and were diagnosed by an experienced neurologist. The diagnosis was based on history, neuroimaging, neurological examination, and neuropsychological testing, and all individuals met the current criteria for MCI. The exclusion criteria for the study included individuals 1) who were younger than 18 years old, 2) who had a lack of English competence, 3) who had significant psychiatric illness and alcohol and drug use, 4) who had significant neurological problems affecting the brain (e.g., stroke, multiple sclerosis, and Parkinson’s disease), and 5) who had uncorrected visual or hearing loss. All individuals with MCI fulfilled the recent criteria of the 2018 National Institute on Aging-Alzheimer’s Association (NIA-AA) research framework [43].

Demographic information for individuals with MCI can be found in Table 1.

**Table 1.**
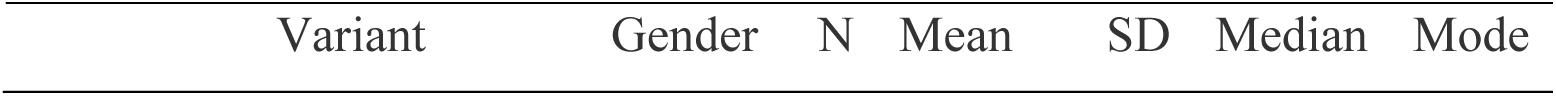
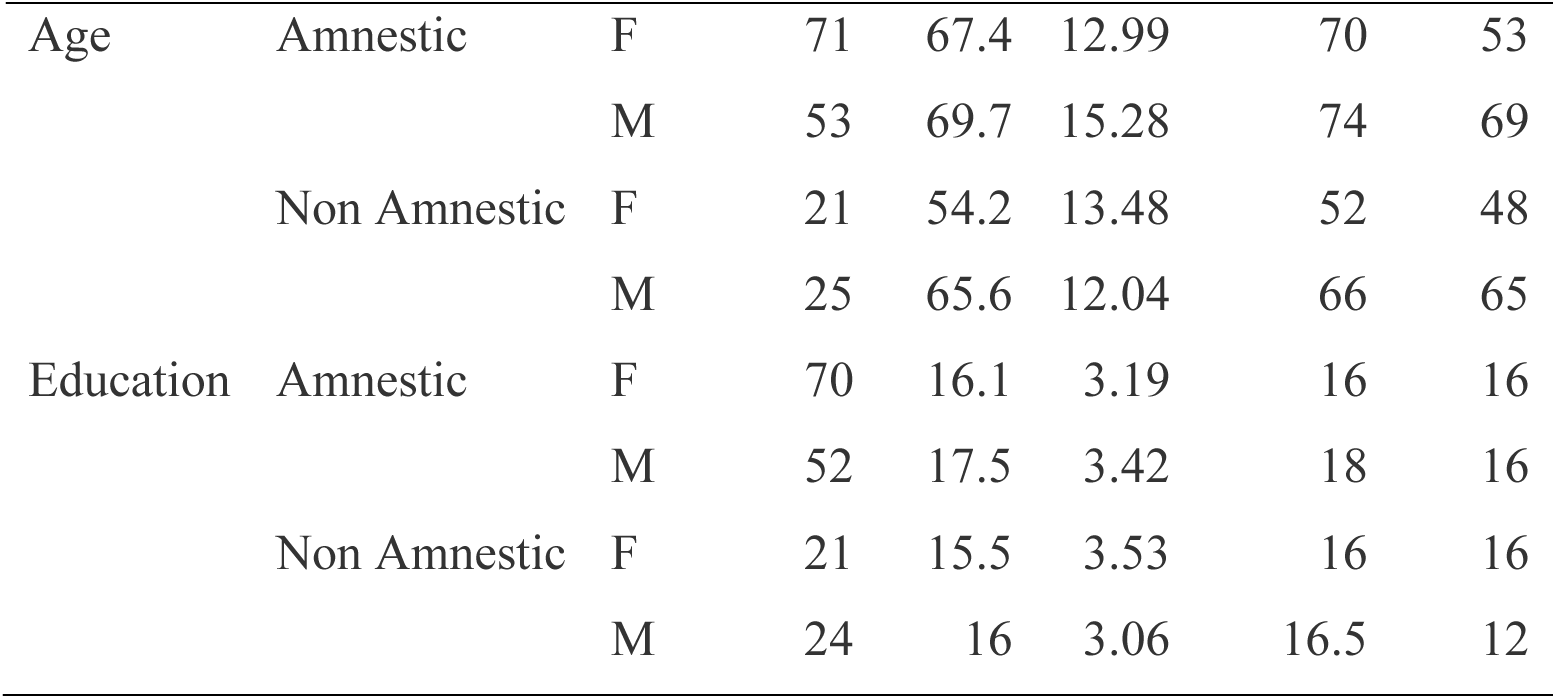
Participants’ Age and Education across variants (Amnestic and Non-Amnestic) and gender.

Specifically, participants underwent a battery of standardized neuropsychological tests to assess their cognitive and linguistic abilities. These tests comprehensively evaluated various aspects of language and cognitive functioning, offering a detailed assessment of their cognitive strengths and weaknesses. The neurocognitive tests include the Mini-Mental State Examination (MMSE, Folstein, Folstein [44]), the Orientation and Information subset from Wechsler Memory Scale-Third Edition (WMS-III; Wechsler [45]), Digit span subtests of the WMS-III [45], Rey Auditory Verbal Learning Test (RAVLT; Rey, 1941), Rey Complex Figure (RCF; Rey [46]), Boston Naming Test [30], Verbal Fluency Task (FAS), the Free narrative writing section from BDAE [30], Trail Making tests (TMT; Reitan and Wolfson [47]), and Stroop test [48]. The tests were carefully selected to provide a sensitive measure of abnormalities compared to individuals with normal cognitive functioning. Table 2 includes neurocognitive test results for all individuals with MCI. The study protocol underwent rigorous review and received approval from the Johns Hopkins Institutional Review Board (IRB00266221). The data were collected between November 1^st^, 2020, and May 30^th^, 2022. They were subsequently accessed on August 1^st^ 2023 for the purposes of this study. The authors had no information to identify the participants.

**Table 2.**
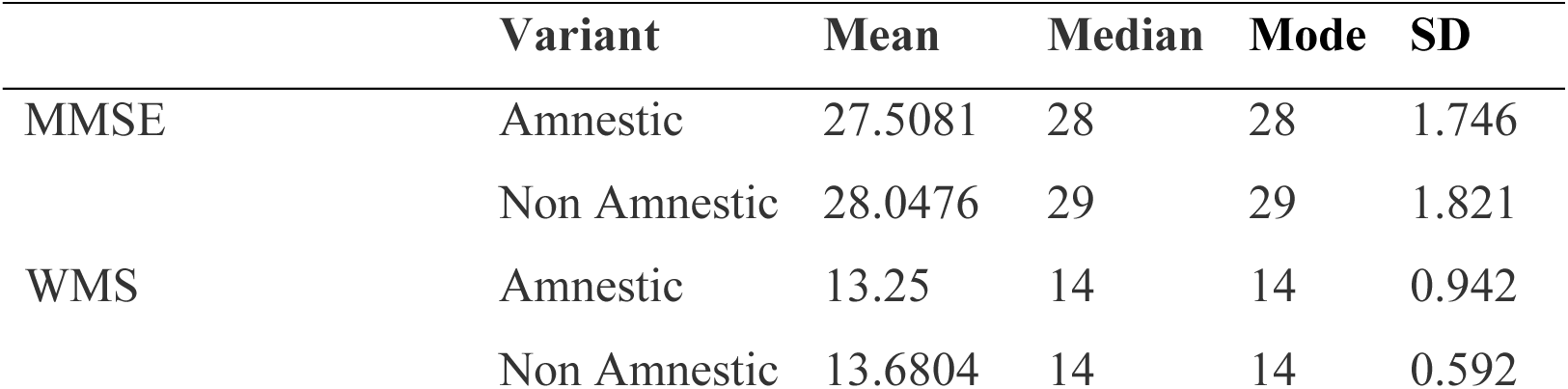
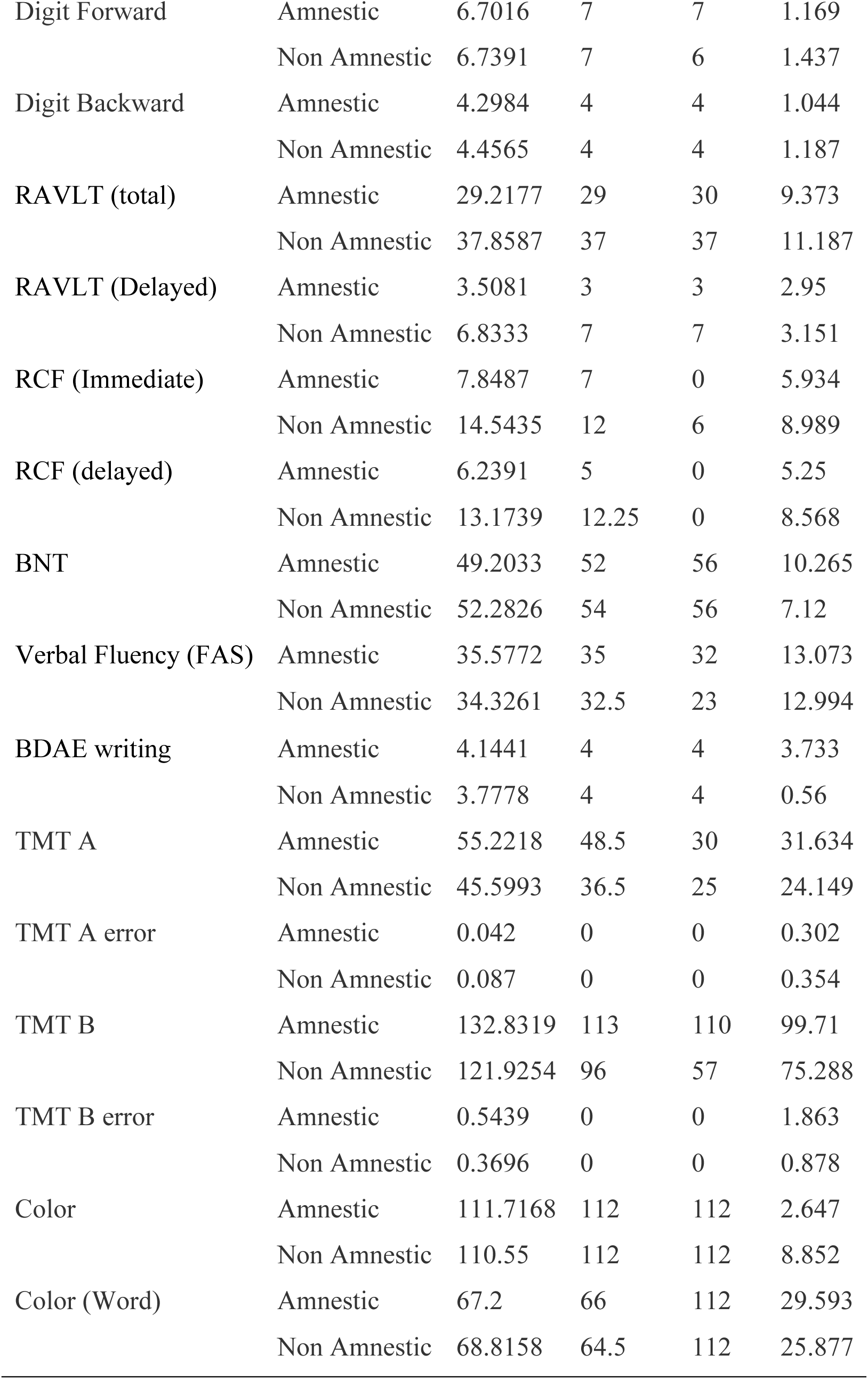
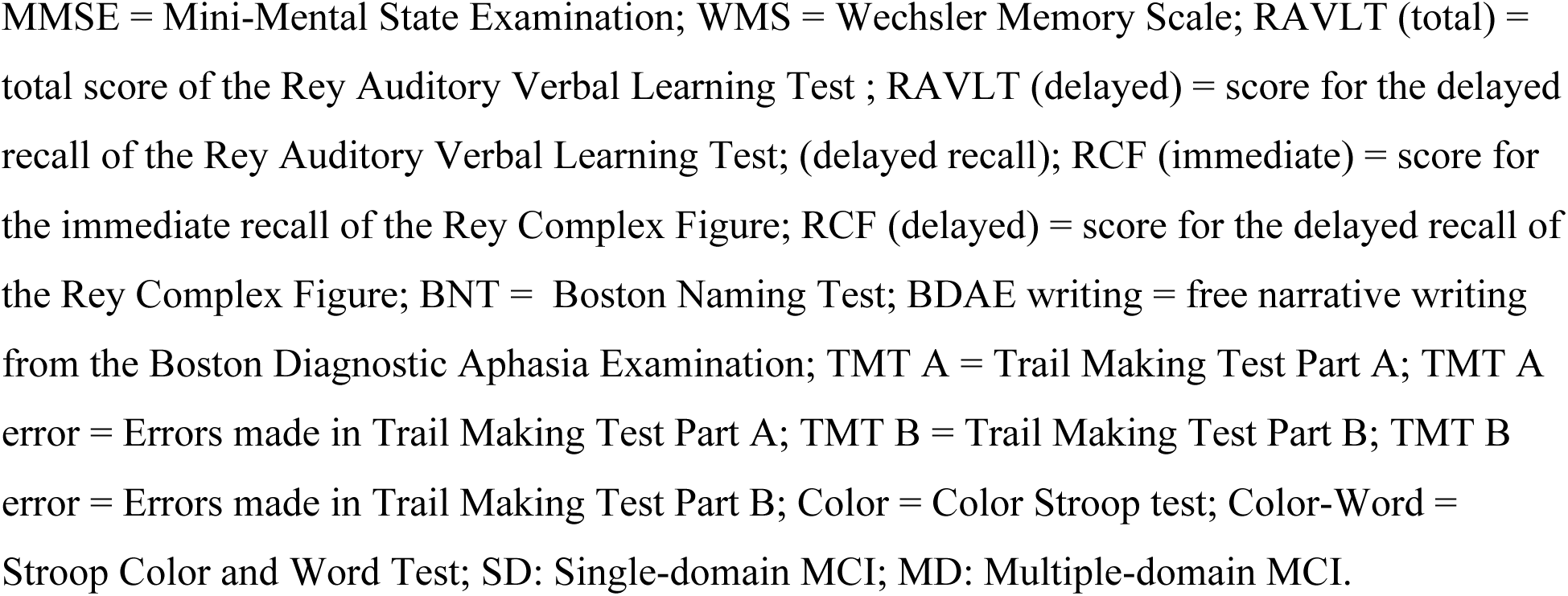
Performance in Neurocognitive Testing in Individuals with MCI.

### 2.2 Written Picture Description Task

Writing samples were collected using the Cookie Theft picture from the Boston Diagnostic Aphasia Examination-3 (BDAE-3; Goodglass, Kaplan [30]. Participants were seated with the picture stimulus and a piece of paper. The clinicians used the prompt to encourage the participants to provide a written description, “Write as much as you can about what you see going on in this picture.” Once the participants completed the task, their writing samples were transcribed into a text document by experienced researchers.

### 2.3 Machine Learning Process

The analysis involved the preprocessing of the data, the extraction of significant features from the written picture description task, and the study of those measures.

#### Analysis of narrative speech

We analyzed the written transcripts from the text documents using two natural language processing tools, including the tokenization of the text, the tagging of morphological categories, and the parsing of the syntactic constituents. Specifically, each word in the text was labelled using Open Brain AI’s POS tagger and syntactic dependency parser, which uses a variety of linguistic information to determine the dependency structure of a sentence [49]. Open Brain AI provided automatic measures that included counts and the ratio of each word / total count of words that appears in the text for each participant.

Specifically, the automatically elicited morphosyntactic measures shown in Table 3 include Part of Speech (POS) categories (i.e., *adjective, adposition, adverb, auxiliary verb, coordinating conjunction, determiner, interjection, noun, numeral, particle, pronoun, proper noun, subordinating conjunction, symbol, verb*), the number of words and characters and their character/word ratio, and syntactic dependency measures indicating the grammatical relationships between words in a sentence and their count to total word ratio.

**Table 3.**
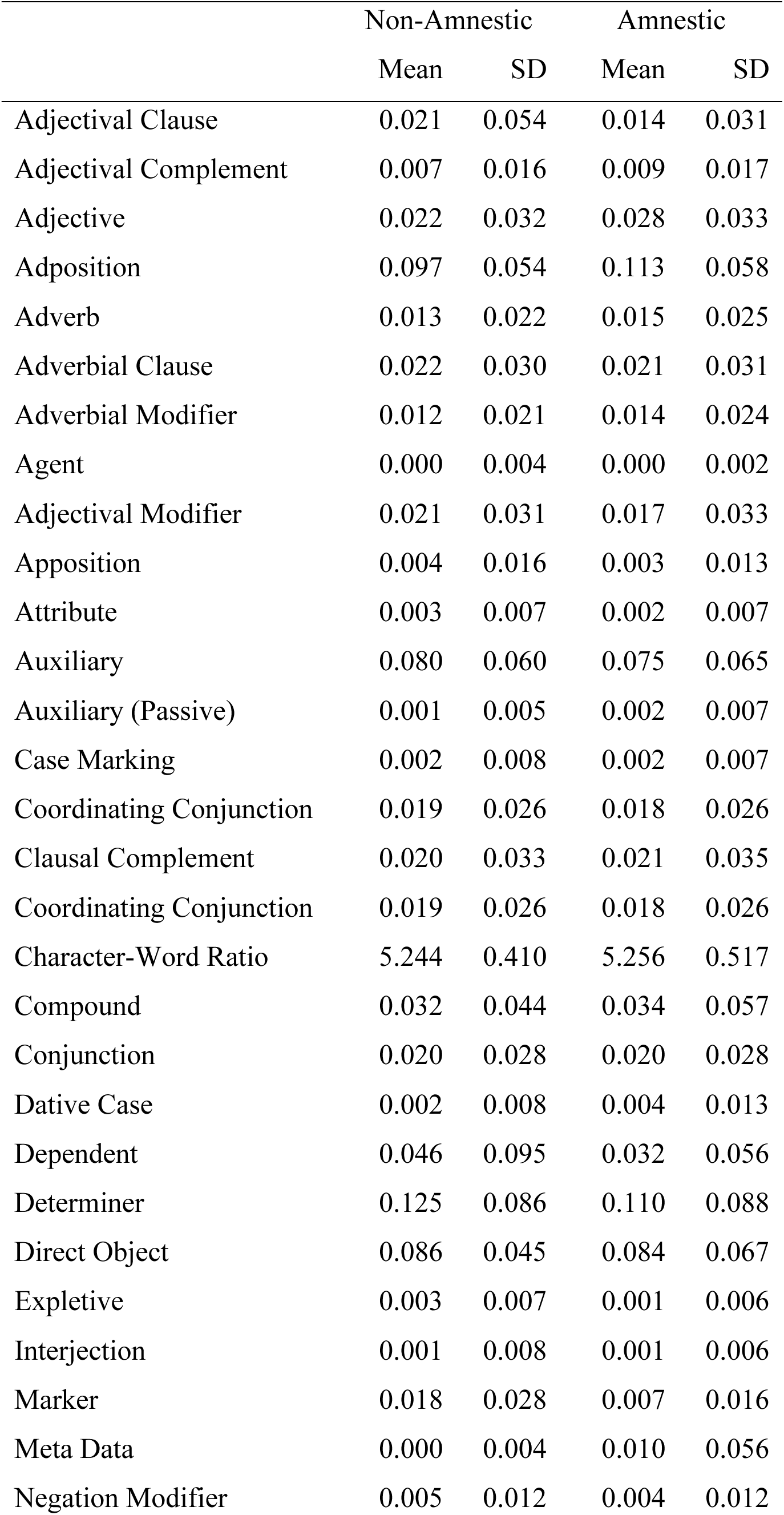
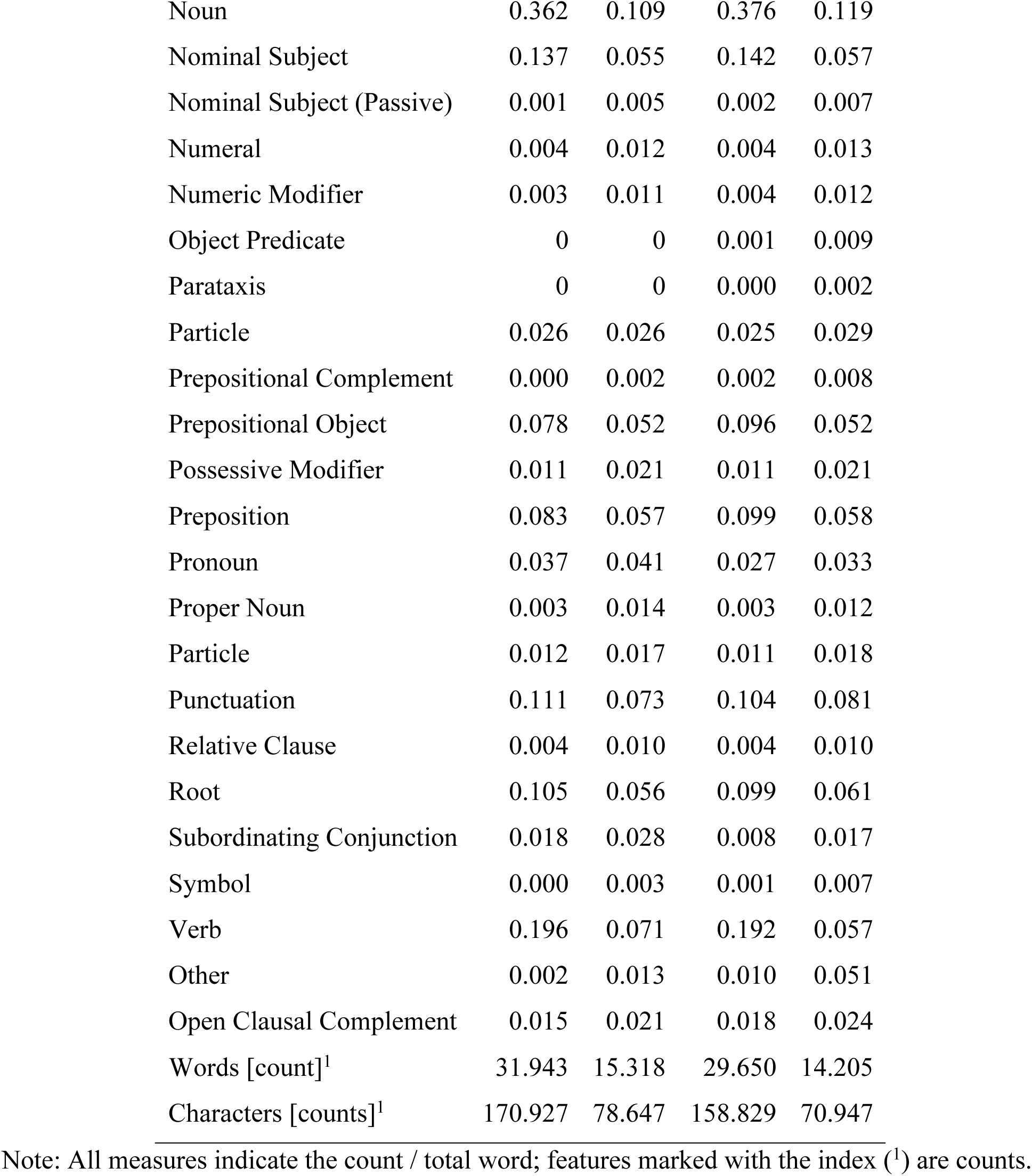
Means and Standard Deviations of features in individuals with Non-Amnestic and Amnestic MCI.

**Table. 4.**
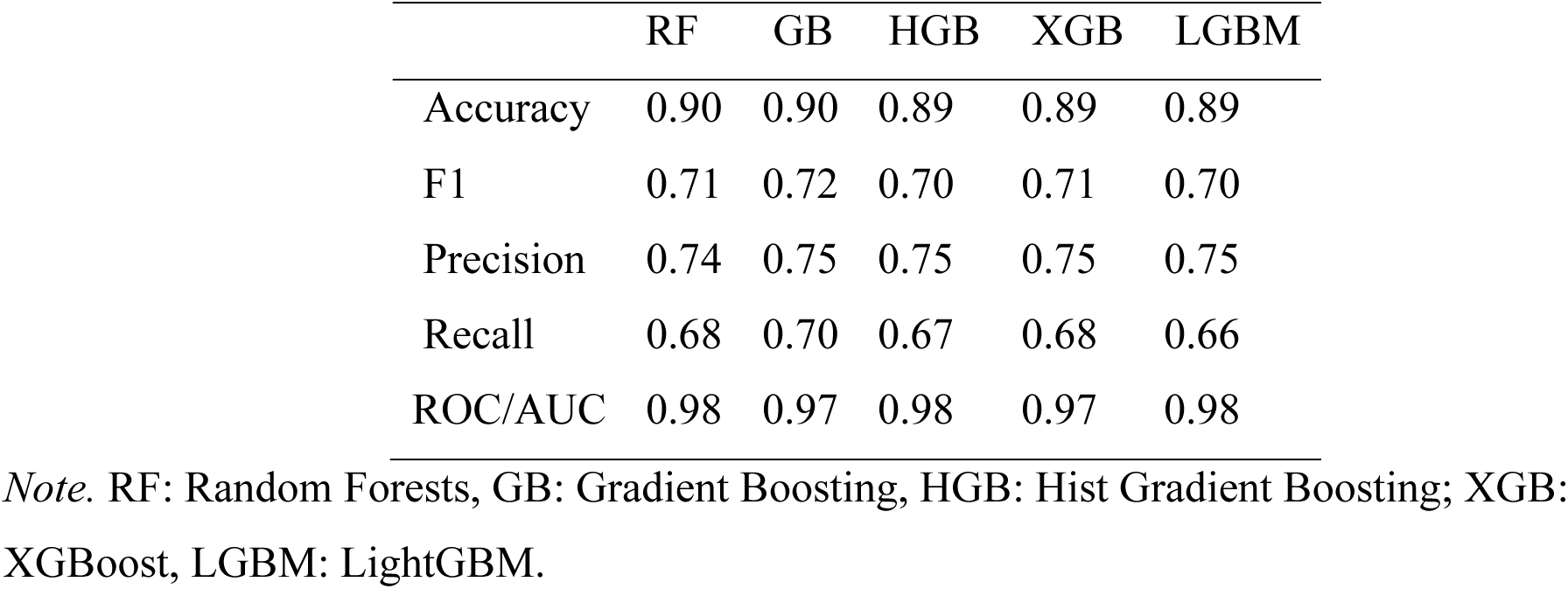
Model performance in the classification task: individuals with aMCI vs. individuals with naMCI from language measures.

#### Semantic measures from BERT

Semantic measures are crucial in individuals with aMCI and can differentiate individuals aMCI and naMCI. To depict semantic relationships, we included word and sentence embeddings from BERT-large-uncased, a BERT (Bidirectional Encoder Representations from Transformers) pretrained language model [50]. Specifically, the BERT-large-uncased is a deep neural network trained on a large dataset of text corpora and can be used for various natural language processing (NLP) tasks, such as question answering, text summarization, and sentiment analysis. The BERT-large-uncased has been shown to achieve state-of-the-art performance on various NLP tasks. It consists of 12 encoder layers, each containing a self-attention mechanism and a feed-forward network. The self-attention mechanism allows the model to learn long-range dependencies between words in a sentence, while the feed-forward network adds non-linearity.

#### 2.3.1 Addressing Imbalance and Cross-validation

We employed Random Over-Sampling (ROS) to balance the class distribution and address the limitations of the relatively small dataset [51]. This technique alleviates the models’ tendency to favor the majority class, a common challenge in imbalanced datasets. Additionally, we implemented group 5-fold cross-validation. This approach minimized data leakage and provided a more reliable model performance evaluation. Furthermore, we standardized the non-BERT features to ensure uniformity in scale.

#### 2.3.2 Model Evaluation and Selection

We selected ML models that do not require massive amounts of training data. To choose the best model for our data, we have trained ML models that roughly belong to four main categories of models, namely ensemble learning models (Random Forest (RF), Gradient Boosting (GB), XGBoost (XGB), and LightGBM (LGBM)). RF is a ML method combining several decision trees to enhance prediction accuracy. This approach can manage high-dimensional data and is resilient to overfitting. GB sequentially combines weak ML learners, each correcting the predecessor’s errors. GB is used in classification and regression tasks for large, complex datasets. XGB and LGBM implement gradient boosting with speed and accuracy. They are employed in scenarios requiring rapid processing of large datasets. Hist Gradient Boosting (HGB), a gradient boosting variant, uses histograms for feature representation, enhancing efficiency with large-scale, high-dimensional data structures. Each ML algorithm has unique strengths, making these models suitable for specific data types and prediction tasks. Only comparing and selecting ML models provides versatility, adaptability, and improved performance in the ML process, enabling the model to tackle the various underlying characteristics of the data.

#### 2.3.3 Hyperparameter Tuning and Model Comparison

A grid search with cross-validation was employed to evaluate and compare the performance of the different machine-learning models. The hyperparameter tuning involved finding the optimal hyperparameters for each model using grid search and calculating the evaluation metrics.

Grid search is a method for hyperparameter tuning evaluates different combinations of predefined hyperparameter values to determine the combination that produces the best performance for a given model. In this case, a grid search was performed for each of the machine learning models included in the study.

We evaluated each model using five-fold cross-validation, which involves evaluating the performance of a model by splitting the data into multiple folds. Each fold is used as a validation set, while the remaining folds are used as the training set. The model is trained on the training set and evaluated on the validation set. This process is repeated for each fold, and the average performance across all folds is used as the final performance estimate.

Various evaluation metrics were used to assess the performance of the different machine learning models. These metrics included accuracy, F1 score, precision, recall, ROC AUC, and Cohen’s kappa score: i. Accuracy is the proportion of correct predictions; ii. F1 score measures a model’s ability to correctly classify positive and negative cases; iii. Precision is the proportion of positive predictions that are positive; iv. Recall is the proportion of positive cases correctly classified as positive, and v. ROC AUC (Receiver Operating Characteristic Area Under the Curve) measures a model’s ability to distinguish between positive and negative cases.

## 3 Results

Written picture description tasks were processed using combined NLP analysis and Bidirectional Encoder Representations from Transformers (BERT) models to elicit measures representing the embeddings. We have implemented two machine learning-supervised classification tasks.

A classification model was designed to distinguish individuals with aMCI and naMCI. The model included only information from the Cookie-Theft picture description task. The model distinguished individuals with aMCI and naMCI. These results suggest that the written discourse from a picture description task provides sufficient information to identify the individuals with the two variants of MCI.

In the ML models, the ROC curves were nearly 98% for classifying individuals with aMCI and naMCI (Figure 2). This suggests that written discourse productions, as manifested in a picture description task, can distinguish the two groups of individuals from language measures. Regarding accuracy, the ensemble models with boosting had the best performance (Table 3).

**Figure 1.**
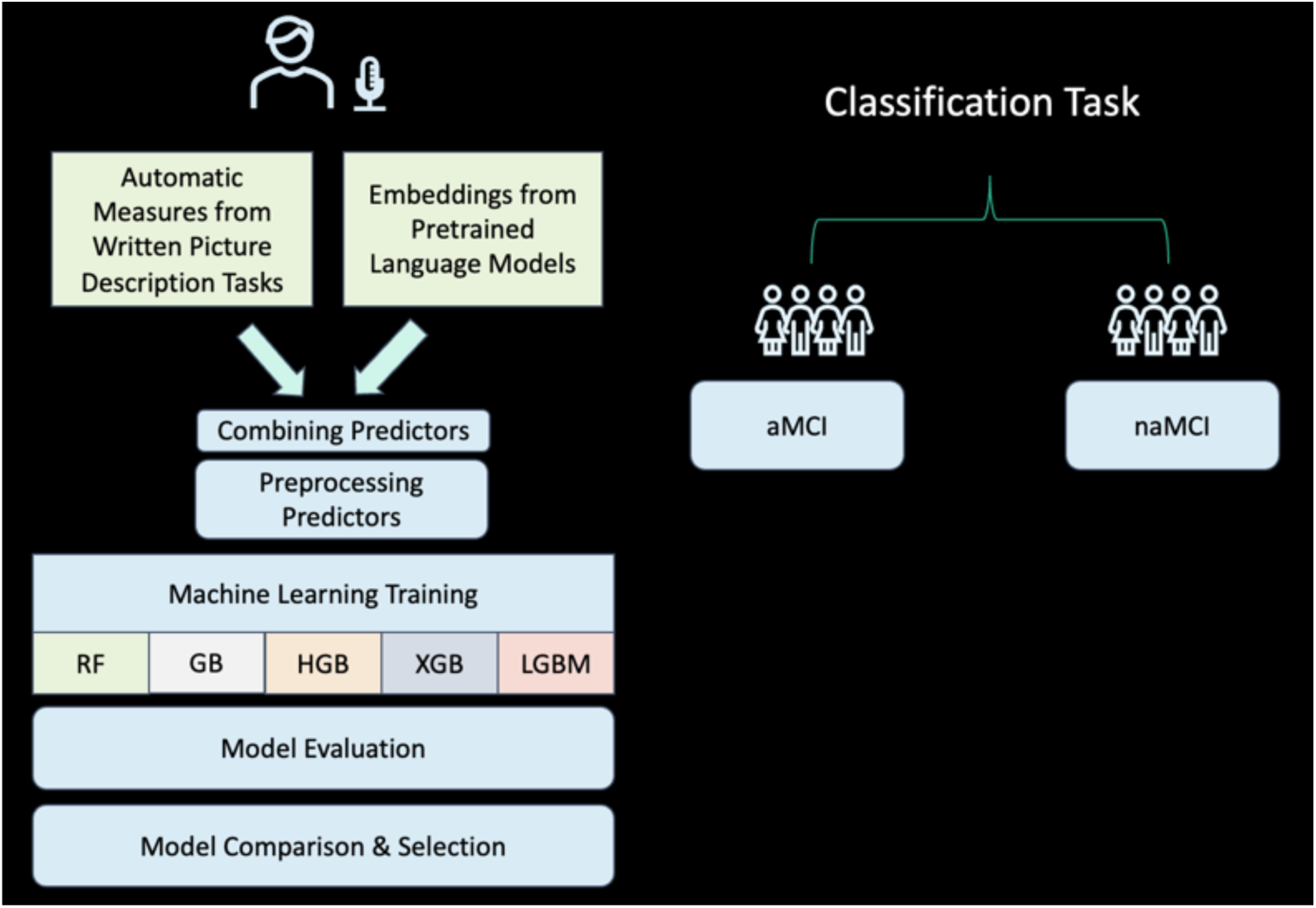
The machine learning classification process and classification task.

**Figure 2.**
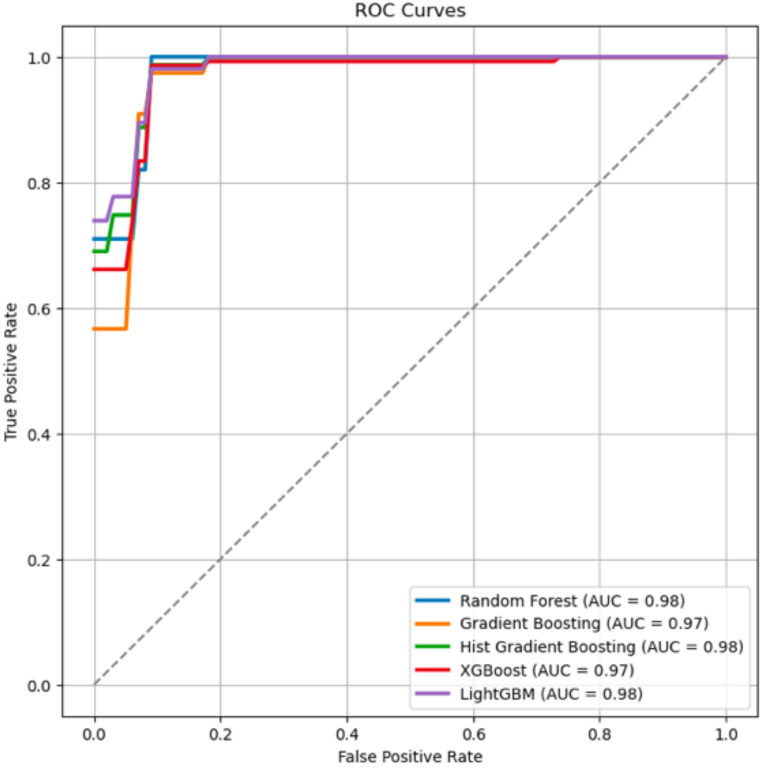
MLs performance on the classification task: individuals with aMCI vs. individuals with naMCI from language measures.

Gradient Boosting, Hist Gradient Boosting, XGBoost, and LightGBM. The consistency in the output of those models further demonstrates their effectiveness for real-world applications.

As indicated by the outcomes, the utilization of machine learning models shows the potential of MLs in diagnosing and differentiating the two MCI subtypes. The reported standardized metrics – Accuracy, F1 Score, Precision, Recall, and ROC/AUC – indicate the effectiveness of these models, with one (1) being the best value.

- Accuracy (0.90 for most models) reflects the ML model’s overall correctness in classifying the MCI type.
- F1 Score balances precision and recall, with values around 0.70-0.72, indicating a good balance between false positives and false negatives.
- Precision (0.74-0.75) measures the proportion of correctly identified positive cases among all positive calls made by the model.
- Recall (ranging from 0.66 to 0.70) indicates the model’s ability to identify all actual positive cases.
- ROC/AUC (between 0.97 and 0.98) reflects the model’s ability to distinguish between the two classes across various thresholds, with values close to 1 indicating excellent performance.

We have evaluated feature importance. BERT features dominate the rankings of the 15 contributing factors for the RF classification. The following features contribute to RF classification, from more important to less important: prepositional object, adposition, dependent, particle, auxiliary, root (verb), adjective, and subordinating conjunction.

These results suggest a reliable performance in distinguishing patients with naMCI vs. aMCI highlight the potential of advanced machine-learning techniques in medical diagnostics, especially for complex conditions like MCI. The high performance of these models suggests that they could be valuable tools in clinical practice for early and accurate identification of MCI types, thereby enabling more tailored and effective treatment strategies.

## 4 Discussion

MCI is an early stage of cognitive decline due to pathology reasons [4]. Individuals with aMCI are characterized primarily by memory deficits, while individuals with naMCI are impaired in other cognitive functions, such as language, attention, and executive functions.

Identifying the type of MCI is important for predicting the progression of the condition, as individuals with aMCI are more prone to progress into Alzheimer’s disease [52, 53] or all types of dementia (Glynn et al., 2021). This study aimed to determine the potential diagnostic utility of computational methods in classifying two subtypes of MCI. We found that a written picture description task can distinguish individuals with aMCI and naMCI at approximately 90% accuracy. This finding confirms that written discourse analysis, which is infrequently done in clinical settings, provides clinically essential information (Kim et al., 2023) and can be a powerful approach for better characterizing the subtypes of MCI.

Importantly, our study shows that a single behavioral task (i.e., a picture description task) can provide substantial information about domains that require multiple separate tasks. As mentioned earlier, either multiple pen-and-pencil tasks or neuroimaging techniques need to be conducted clinically to classify MCI. Previous studies using machine learning algorithms and neuroimaging data demonstrated an accurate classification of MCI subtypes [54, 55]. However, data can be obtained only with advanced techniques. They are not often feasible for individual patients [56]. Behaviorally, multiple tasks that evaluate different cognitive components, such as memory and executive functions, need to be administered, which is considered a time-intensive process. From a clinical perspective, computational assessment of language with machine learning and natural language processing opens the door for exciting opportunities to expand the analysis to both longer and more complex tests productions.

Besides the cost-effective assessment, it is also significant to note that the current study used written discourse samples, which received little attention in research (Kim et al., 2023) and are not often collected and evaluated in clinical settings [57]. He, Chapin [58] used a spoken discourse task to investigate the classification among healthy adults, subtypes of MCI, and dementia. In the study, the researchers used both linguistic and acoustic features, but the classification accuracy (aMCI vs naMCI) was 88%. Our findings shed light on the clinical value of written discourse as the linguistic features in writing lead to higher classification accuracy.

This also indicates that linguistic features in writing can be potential markers of memory deficits and may provide enough information for the classification.

Written discourse offers a plethora of information about individuals’ linguistic functioning, including textual macrostructure and microstructure. However, it is not clear which components of written discourse in this population are more influenced by cognitive impairment in MCI. This is evidenced by 102 different measures used to quantify writing behaviors in research with little repetition of the same measure (Kim et al., 2023). In the current study, using written discourse samples, we calculated the POS of each word and syntactic relationships [59] that appears in the written picture description task [60]. Together, this can be an optimal approach for analyzing such language samples in that it adds to the efficiency of written picture description analysis. This also provides a comprehensive and detailed grammar analysis in a standardized and less subjective manner.

Moreover, we found that the BERT semantic features dominated the hierarchy of analytical constructs that we have used. This finding is consistent with the consensus that impairments in semantic domains of language are a key manifestation of disease progress in neurodegenerative disorders [61–63]. These features can be seen in the literature to be associated with one or more elements of the writing skills of individuals with MCI, as they interface linguistic and semantic memory domains.

Specifically, context-sensitive embeddings from BERT [50] played a critical role in the high accuracy of the classification. These result from averaging the token-level embeddings from the last layer of a BERT model for each input text, which creates a single, comprehensive vector representation for the entire text, capturing its overall contextual meaning. Traditional word embedding techniques, such as Word2Vec [64] and GloVe [65], generate a single word embedding for each word in the vocabulary. The embeddings are decontextualized, which fails to capture the meanings of polysemous words. For instance, the word *bank* can mean a financial institution that accepts deposits and makes loans or the sloping edge of a river or other body of water. On the other hand, BERT uses a technique known as contextual embedding. This means that the representation of a word is based on sentence context. So, the word *bank* would have different representations in the sentences “*I went to the bank to retrieve money”* and “*the little house next to the river bank,”* which offers a better representation of ambiguous meanings, improving the accuracy of text classification. Again, these contextual embeddings utilized in this study demonstrate a better understanding of the syntactic and semantic relationships between words in a sentence. This is crucial for quantifying the overall thematic content of the written picture descriptions. Additionally, since individuals with amnestic and non-amnestic MCI differ in their semantic memory [66, 67], the contextual sensitivity of BERT’s embeddings helps the model to adapt to differences in vocabulary and jargon.

Although it is well-known that picture description tasks are valuable for eliciting connected language samples in individuals with MCI [68], the Cookie-Theft picture offers a less ecological way of personal expression through writing. Such productions are constrained substantially in their context and effectively identifying differences in pragmatic language usage and speech and voice parameters. Also, the task does not allow the assessment of non-epistemic domains, such as deontic modality expressions of wish and hope and non-present tense verb-tense semantics, as it does not provide opportunities to discuss past or future events.

Additionally, picture description tasks do not offer opportunities for expressing emotional and other affective content, which might be necessary for assessing the interface of language, emotion, and pragmatics. An open-ended essay writing could have offered the potential to assess more stylistic, linguistic, and communicative speech characteristics. Nevertheless, written picture descriptions demonstrate the potential to detect speech and language characteristics in neurodegenerative diseases such as MCI and dementia, as suggested by a recent review (Kim et al., 2023). Considering the brief time to elicit writing samples, NLP combined with discourse-level writing samples will enable more efficient methods for analyzing these linguistic and communicative features, further enhancing the diagnostic accuracy and the clinical utility of written discourse analysis.

The results of the current study suggest that written discourse samples can offer a quick and efficient means of gaining valuable insights into linguistic abilities while minimizing the burden placed on individuals with MCI. Future research is necessary to verify this finding with a balanced sample size between aMCI and naMCI. For a better diagnostic tool, future studies, including MCI-dementia conversion, are needed to test the predictive value of the automatic classification of MCI.

## Data Availability

Data are available upon requests from the authors.

## Conflicts

HK, AH, and CT do not have any conflicts of interest.

## Funding Sources

No funding source available.

## Consent Statement

All human subjects provided informed consent.

## Notes

### Competing Interest Statement

The authors have declared no competing interest.

### Funding Statement

The author(s) received no specific funding for this work.

### Author Declarations

IRB of Johns Hopkins University gave ethical approval for this work (IRB00266221). The data were collected between November 1st, 2020, and May 30th, 2022. They were subsequently accessed on August 1st 2023 for the purposes of this study.

### Summary of Updates

This version now corresponds to the one submitted to a journal and includes various grammar and style updates and the complete author specification.

